# Effect of Urinary tract infection on the outcome of the Allograft in patients with Renal transplantation

**DOI:** 10.1101/2024.02.06.24302324

**Authors:** Rahul Sai Gangula, Mahesh Eswarappa, Rajashekar Reddy, Gireesh Mathihally Siddaiah, Gurudev Konana, Hamsa Reddy, Pooja Prakash Prabhu, Yousuff Mohammad, Lia Sara Anish

**Affiliations:** Department of Nephrology, M. S. Ramaiah Medical College, Bengaluru, India

**Keywords:** Urinary tract Infection, Recurrent UTI, Non-Recurrent UTI, Transplantation

## Abstract

**Background:** Urinary Tract Infections (UTIs) are the second most common cause of graft dysfunction, accounting for significant morbidity, and are associated with poor graft and patient survival. This study aimed to determine the association between post-renal transplant UTI and graft outcomes.

**Methods:** We examined the effect of UTIs on graft outcomes in patients who underwent renal transplantation surgery between January 2010 and December 2022. The study population included 349 renal transplantations, of which 74 experienced 140 UTI events. Based on the number of UTI episodes, patients were categorized into three groups

**Results:** Of the 349 recipients, 275 (74.4%) had no UTI, 47 (18.8%) had nonrecurrent UTIs (NR-UTIs), and 27 (6.8%) had Recurrent UTIs (R-UTIs). NR-UTIs were associated with very poor graft survival compared with no UTI (Hazard Ratio [HR], 2.312; 95% Confidence Interval [CI], 1.410–3.791; P=0.001). This relationship persisted even after adjusting for confounding factors in Multivariable Cox regression analysis (HR, 2.318; 95% CI, 1.414–3.800; P=0.001). Although R-UTIs appeared to result in poor patient survival, the difference was not significant (vs No UTI, HR, 1.517; 95% CI, 0.983–2.342; P=0.060). There appeared to be higher patient survival in R-UTIs but was not significant (vs NR-UTI, HR, 1.316; 95% CI, 0.486–3.564; P=0.589). R-UTIs were more likely to be associated with Multi-drug Resistant Gram-negative organisms (Klebsiella pneumonia or Escherichia coli) with resistance to Nitrofurantoin (RR, 2.753; 95% CI, 1.257–6.032; P=0.01) and Carbapenem (RR, 2.064; 95% CI, 0.988–4.314; P=0.05).

**Conclusion:** NR-UTIs were associated with poorer graft and patient outcomes than no UTI.

## Introduction

End-stage renal disease is the terminal stage of chronic kidney disease, in which the kidneys can no longer support the body’s needs. Although various modalities of renal replacement therapies are available, kidney transplantation ensures a maximum life span with the best quality of life and is the most cost-effective. Hence, kidney transplantation is currently considered to be the best modality for Renal Replacement therapy.

After cardiovascular disease, infections are the second most common cause of mortality in renal transplant recipients^1^. Urinary Tract Infections (UTIs) are among the most common infections after renal transplantation and, can lead to graft dysfunction, and compromise the function of the transplanted kidney^1–2^. Although UTIs may occur at any time after renal transplantation, they are most common in the initial year post-transplantation and may lead to sepsis, acute cellular rejection, impaired allograft function, graft loss, and patient death^1–3^. Hence most centers prescribe at least 3–6 months of anti-microbial prophylaxis after renal transplantation, although the regimens may vary^1–4^. Despite routine administration of antimicrobial prophylaxis during the initial post-transplant period, it accounts for significant morbidity and mortality in transplant recipients^4^.

The incidence of UTI in renal transplant recipients is highly variable between studies (7% - 80%). This varied incidence is due to the variability in patient populations, study designs, and definitions of UTIs. Some UTIs may be asymptomatic or cause only mild symptoms, whereas others may lead to severe complications that affect graft function. Approximately 19% of patients develop acute pyelonephritis in the first 2 years after renal transplantation. Common risk factors include prolonged indwelling catheters and Double J stents after transplantation, premature discontinuation of antibiotics, short length of transplant ureter, absence of sphincter between transplant ureter and native bladder, and net high dose of immunosuppression immediately post-transplant^5–7^. Risk factors for recurrence include older age, female donor, deceased donor, neurogenic bladder, history of preoperative UTI, and acute rejection episodes (treated with steroids or immunosuppressive regimens)^5–7^. Recurrent UTIs have been reported to occur in 4%-72% of transplant recipients^8^.

Although most of the studies^8–12^ have shown an association between post-renal transplant UTI and deterioration of graft function, the impact of UTI on long-term graft and patient outcomes is less clear with divergent results. This study was organized with the following objectives to address these critical knowledge gaps:

1. To determine the association between post-renal transplant UTI and graft outcomes.
2. To determine the association between post-renal transplant UTI and patient survival.
3. To describe and compare the microbiological and antimicrobial resistance profiles of patients with Non-recurrent UTIs (NR-UTI) and Recurrent UTIs (R-UTI).

## Materials and Methods

### Study design and patient population

This hospital-based, observational, cohort study was conducted in a tertiary care hospital in Bengaluru, Karnataka, India. The study was approved by the Institutional Ethics Committee (DRP/IFP1085/2023). This study included all the patients who underwent kidney transplantation at our institute between January 2010 and December 2022.

### Definitions and grouping of UTI events

UTI was defined as the presence of any of the following clinical symptoms of fever, dysuria, burning micturition, abdominal or loin pain, foul-smelling urine, increased frequency of micturition, and a urine culture sample revealing single microorganism growth with > 10^5^ bacterial colony-forming units per mL.

All patients were categorized into 3 groups based on their UTI status after renal transplantation (No UTI, Non-Recurrent UTI, and Recurrent UTI).

Recurrent UTI was defined as a patient who had 2 or more UTI episodes in any 6 months (or) 3 or more episodes in any 12 months during post-transplant follow-up.

Non-recurrent UTI was defined as all patients with a history of UTI after renal transplantation who were not classified into the Recurrent UTI group.

No UTI was defined as all the patients who had never experienced any episode of UTI during post-renal transplant follow-up.

Antibiotic resistance was defined according to the Clinical and Laboratory Standards Institute (CLSI) guidelines for antimicrobial susceptibility testing.

Multi-drug resistance was defined as non-susceptibility to at least one agent in three or more different antimicrobial categories.

Graft failure was defined as impaired functioning of the graft kidney in the recipient, requiring renal replacement therapy for more than 3 months.

Asymptomatic bacteriuria was defined as patients who had no symptoms suggestive of a UTI but urine culture had grown organisms with >10^5^ colony-forming units per ml

Acute cystitis was defined as a urinary tract infection confined to the bladder in an otherwise healthy, premenopausal, non-pregnant female.

### Posttransplant immunosuppression

All the patients were stratified according to the need for induction based on dialysis vintage, HLA mismatches, blood group compatibility, recipient age, and donor age. All high-risk patients received ATG, whereas the moderate-risk patients received Basiliximab. The low-risk patients were not administered ATG or Basiliximab. Additionally, all patients received parenteral steroids on the day of transplantation and 2 days post-transplantation. All transplant recipients underwent ‘Lich-Gregoir’ ureterovesical anastomosis.

All the patients received a triple-drug maintenance immunosuppressive regimen consisting of Tacrolimus, Mycophenolate Mofetil, and Prednisolone. Tacrolimus was started at 0.08-0.1 mg/kg/day and subsequently, the dose was adjusted according to Tacrolimus trough levels. Tacrolimus trough target level of 9-10 ng/ml up to 3 months post-transplant, a target of 7-9 ng/ml from 3 to 12 months post-transplant, and a target of 5-7 ng/ml thereafter were considered. Mycophenolate mofetil 500mg twice to thrice daily was administered. Steroids were tapered to 10 mg 3 months post-transplantation and 10 mg was continued until 12 months post-transplantation. Thereafter steroids were tapered to 5-7.5mg per day, which was continued thereafter.

### Catheter and DJ stent policy

A double J-stent was inserted into all allograft recipients during transplantation as a standard procedure and was removed aseptically between 3 and 8 weeks after transplantation. All transplant recipients were placed on Foley catheters during the renal transplantation. They were usually removed between postoperative days 5 and 9 unless the patient had a neurogenic bladder or any other indication where prolonged Foley catheterization was recommended.

### Post-transplant chemoprophylaxis

CMV seronegative patients who received a kidney from a CMV seropositive donor and deceased donor renal transplant recipients were administered valganciclovir during the first 6 months post-transplantation. Additionally, trimethoprim-sulfamethoxazole for Pneumocystis jiroveci was administered during the first 3 months post-transplant in whom it was tolerated.

### Asymptomatic bacteriuria

If a patient was found to have asymptomatic bacteriuria in the initial 2 months post-transplant, urine culture was repeated. If the patient had two consecutive urine cultures that yielded >10^5^ colony-forming units of the same pathogen, they were treated with antibiotics for 5 days according to culture sensitivity. Asymptomatic bacteriuria beyond 2 months post-transplantation was not treated with antibiotics.

### Treatment of UTI

All UTI events without sepsis were treated empirically with third-generation cephalosporins. Events of urosepsis were treated empirically with carbapenems. Following urine culture reports, the prescription was adjusted and appropriate antibiotics were administered for an optimal duration (i.e., 14-21 days). All patients with recurrent UTI underwent thorough clinical, laboratory, and imaging workups to identify the cause and were treated accordingly.

### Post-UTI antibiotic prophylaxis

Transplant recipients with recurrent UTIs were treated appropriately and antibiotic prophylaxis was administered for a duration ranging between 6 weeks - 3 months.

### Diagnosis of rejection, BK virus nephropathy

Rejection episodes were diagnosed using graft kidney biopsy, graded according to the Banff classification^13^, and treated accordingly. PCR screening was performed periodically in all transplant recipients. BK viral nephropathy was confirmed using allograft biopsy tissue SV40 staining.

### Outcomes

The primary outcome was the overall graft survival in patients with UTI. The secondary outcomes included yearly graft function, death-censored graft survival, patient survival, and association between UTI status and antibiotic resistance. Patients who had functioning grafts on the day of death were censored to analyze death-censored graft survival.

### Data collection

Data were collected using a predesigned proforma and subsequently entered into Microsoft Office Professional Plus Excel 2016, Version 16.0 (Microsoft Corp, Redmond, USA). To avoid any possible error, the data entry was cross-checked at two levels (entry into the proforma, and entry from the proforma to the Excel sheet) by two independent observers.

### Statistical analysis

Descriptive statistics were calculated for categorical and continuous variables. The Shapiro-Wilk test of normality was used to check for the type of distribution. A p-value of < 0.05 in the Shapiro-Wilk test of normality was considered significant and the distribution was taken as a non-Gaussian distribution. Quantitative variables with a Gaussian distribution were summarized as means and standard deviations. The quantitative variables, which had a skewed distribution, were summarized as medians and interquartile ranges. All patients were categorized into three groups (No UTI, Non-recurrent UTI, Recurrent UTI) based on post-renal transplant UTI. Renal transplant recipient, donor, and transplantation characteristics were compared using the Chi-square test for categorical data and One-way analysis of variance (ANOVA) or the Kruskal-Wallis H test for continuous variables. Patient, graft, and death-censored graft survival outcomes were computed and compared using Kaplan-Meier Survival analyses (log-rank test, p < 0.05 is considered significant) and univariable Cox regression. The mean and Median survival times with 95% confidence intervals were computed for the graft and patient outcomes.

Cox Proportional Hazard models were derived using variables selected using a backward stepwise approach. Variables associated with graft failure (P < 0.20) were considered for inclusion in the model. Only the variables that significantly altered the relationship between post-transplant UTI status and outcome, resulting in a ≥ 10% change in the associated HR, were included in the final multivariate model. UTI was considered a time-dependent variable. The unadjusted risk ratios for antibiotic resistance were compared with the UTI status for Gram-negative species at each episode and at the patient level, p-value of <0.05 is considered statistically significant. All Statistical analyses were performed using the Statistical Package for the Social Sciences (SPSS) Statistics, Version 25 (IBM Co., Armonk, NY, USA), and DATA *tab* Statistics calculator (Graz, Austria).

## Results

A total of 349 patients underwent renal transplantation during the specified period (2010-2022) and all patients were included in the final analysis (Figure 1). The median follow-up duration was 70 months (IQR 31.5-104 months). A total of 287 patients were male and 62 were female (Figure 2). The mean age of the transplant recipients was 37.49 years with a standard deviation of 11.92 years. During the study period, 74 patients experienced 140 UTI episodes. Of these, 47 (13%) patients experienced NR-UTIs, whereas 27 (8%) patients had R-UTIs. A total of 56(40%) and 84(60%) UTI episodes were noted in the patients with NR-UTI and R-UTI, respectively. The other baseline characteristics are summarized in Table 1.

**Figure 1.**
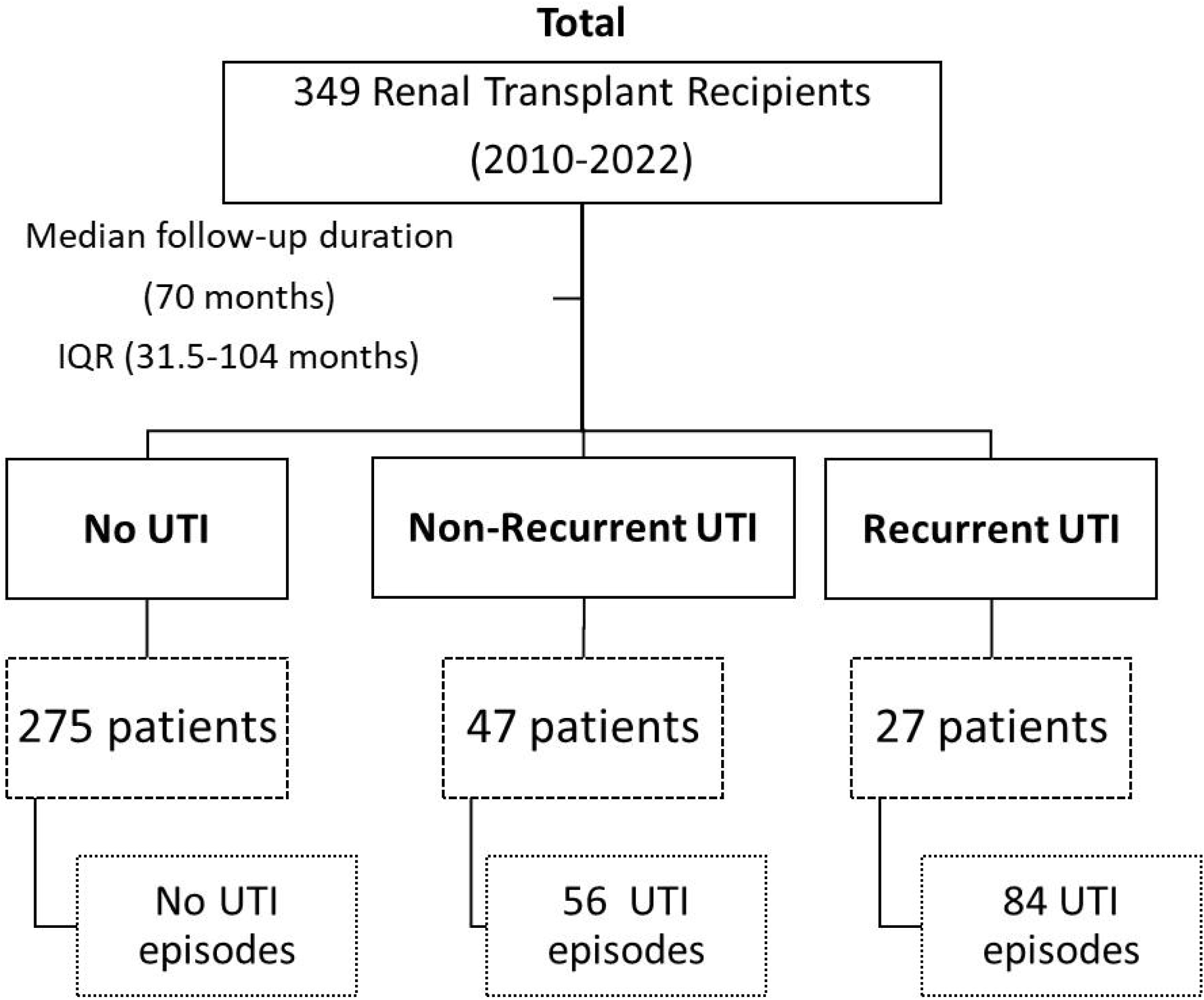
Baseline Flow chart of study design.

**Figure 2.**
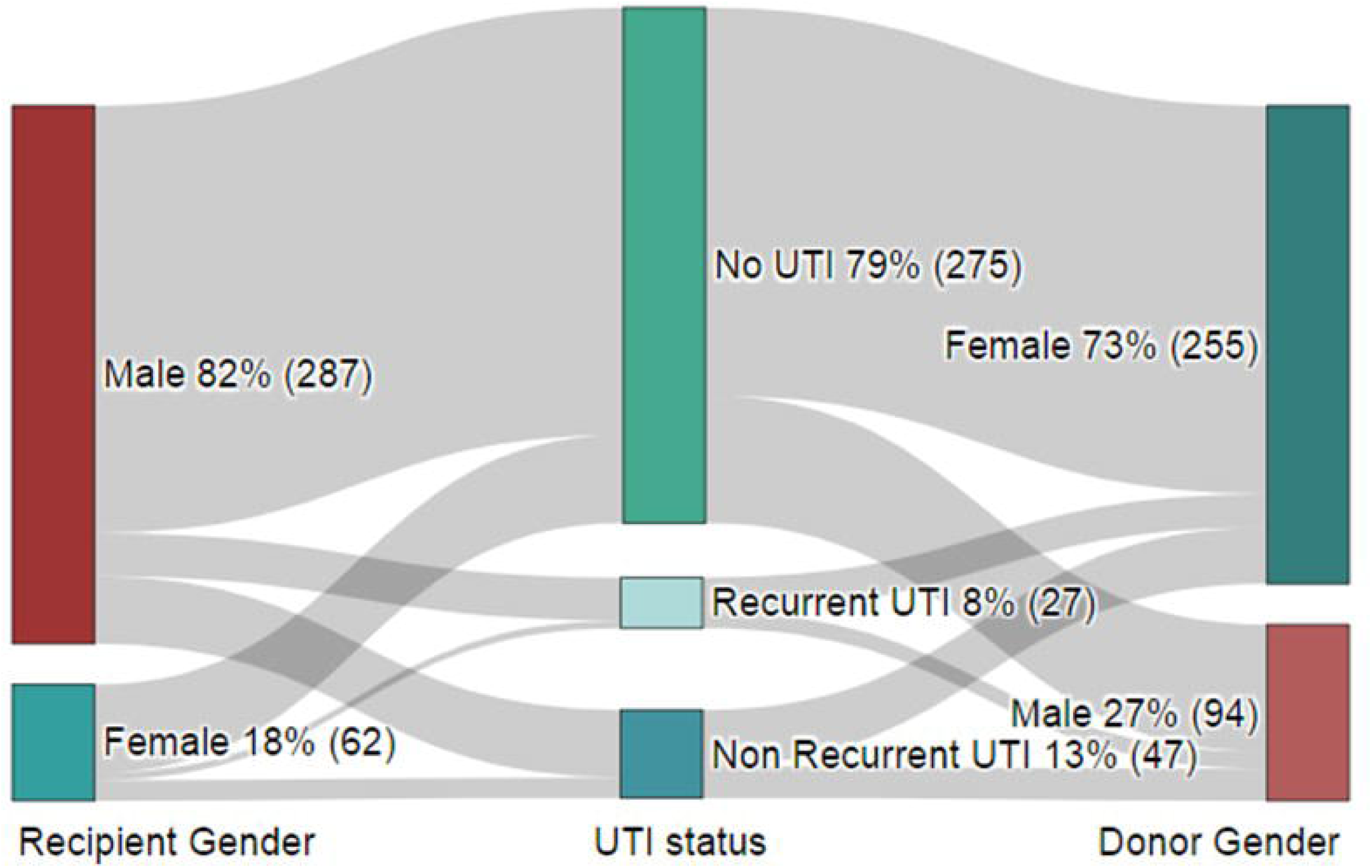
Sankey chart showing the gender of renal transplant recipients and donors.

**Table 1.**
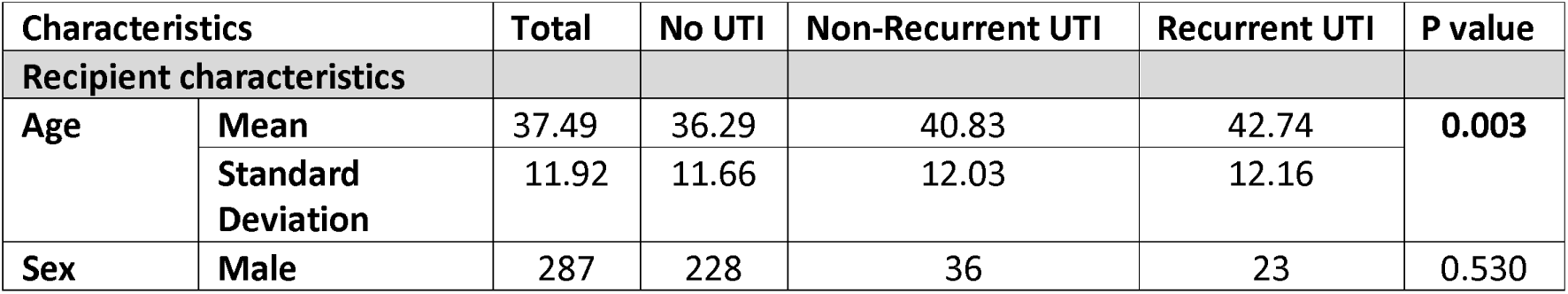
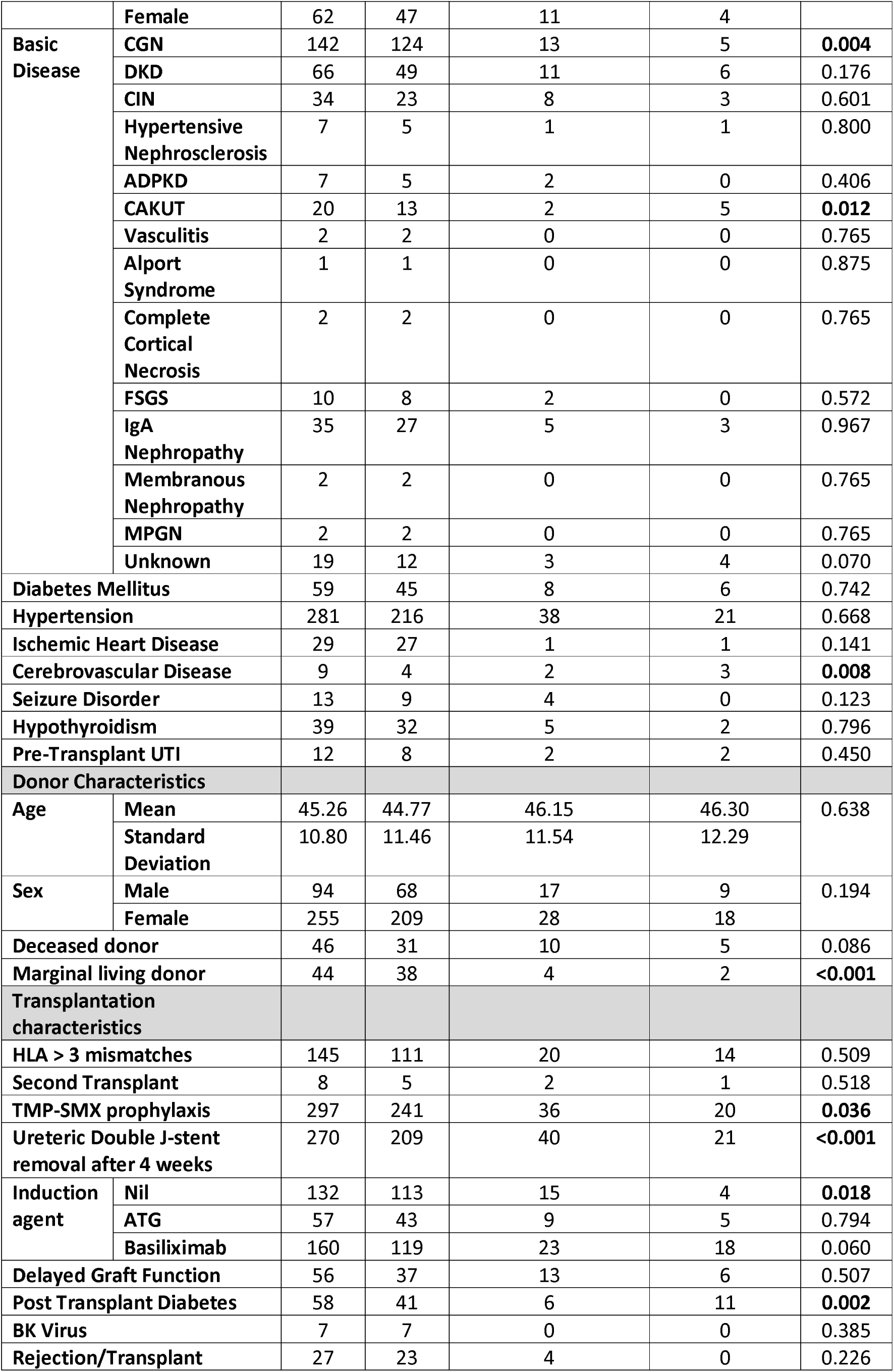
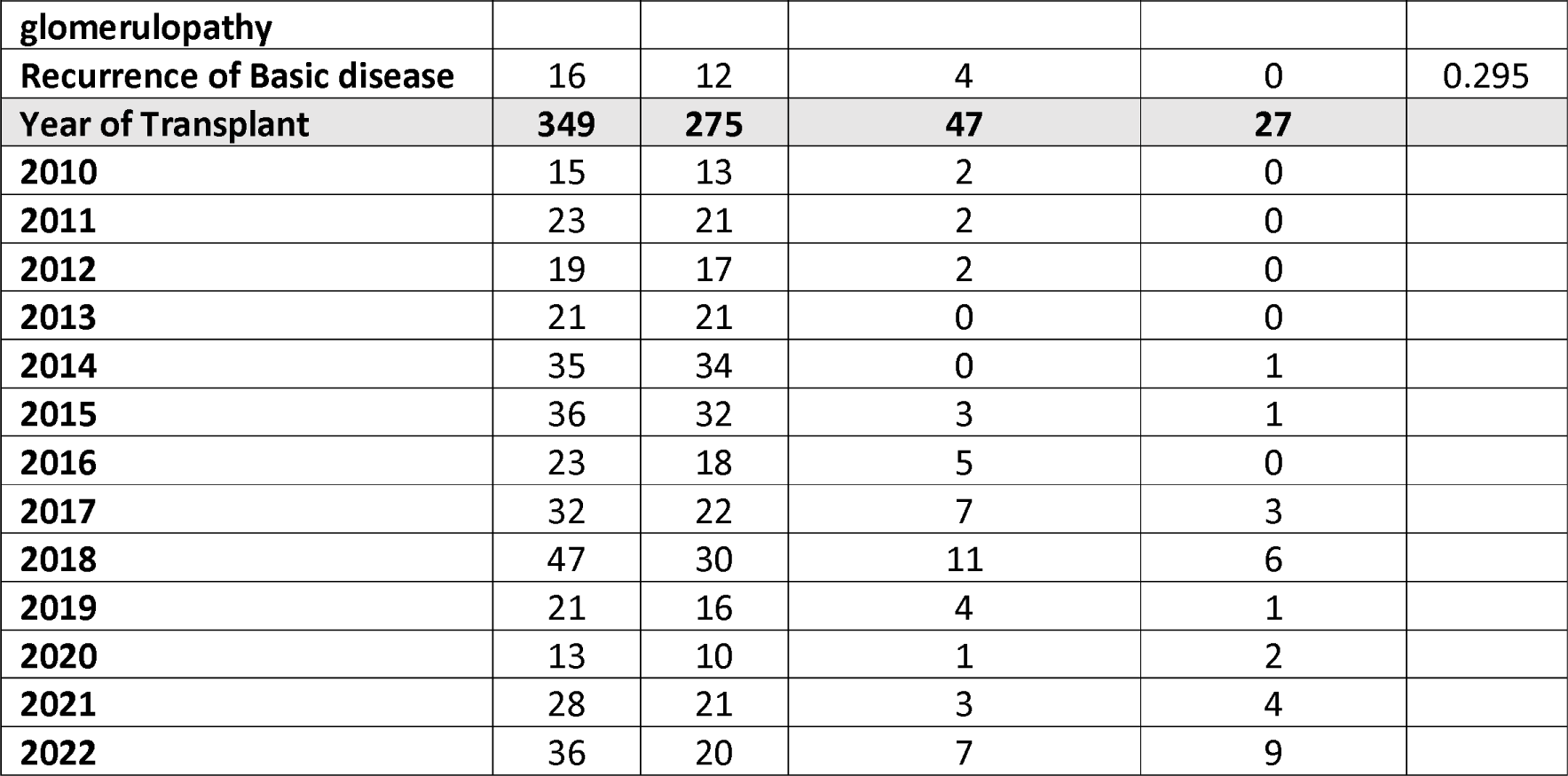
Comparison of Recipient, Donor, and Transplantation-related characteristics based on post-kidney transplant UTI status.

### Recipient characteristics

Recipients with a higher mean age (Figure 3) were more likely to experience NR-UTIs and R-UTI than patients with No UTI events (p=0.039, p=0.019 respectively), and there was no statistically significant difference between NR-UTIs and R-UTI (p=0.779). Recipients with R-UTIs were more likely to have Cerebrovascular disease than patients with No UTI events (p=0.007). However, there was no statistically significant difference between NR-UTIs and patients with No UTI events (p=0.498). Renal transplant recipients with Chronic glomerulonephritis and CAKUT as native kidney disease (Table 1, Figure 4) were associated with R-UTIs (vs No UTI, p=0.019; vs No UTI and NR-UTI, p=0.009 and p=0.029 respectively).

**Figure 3.**
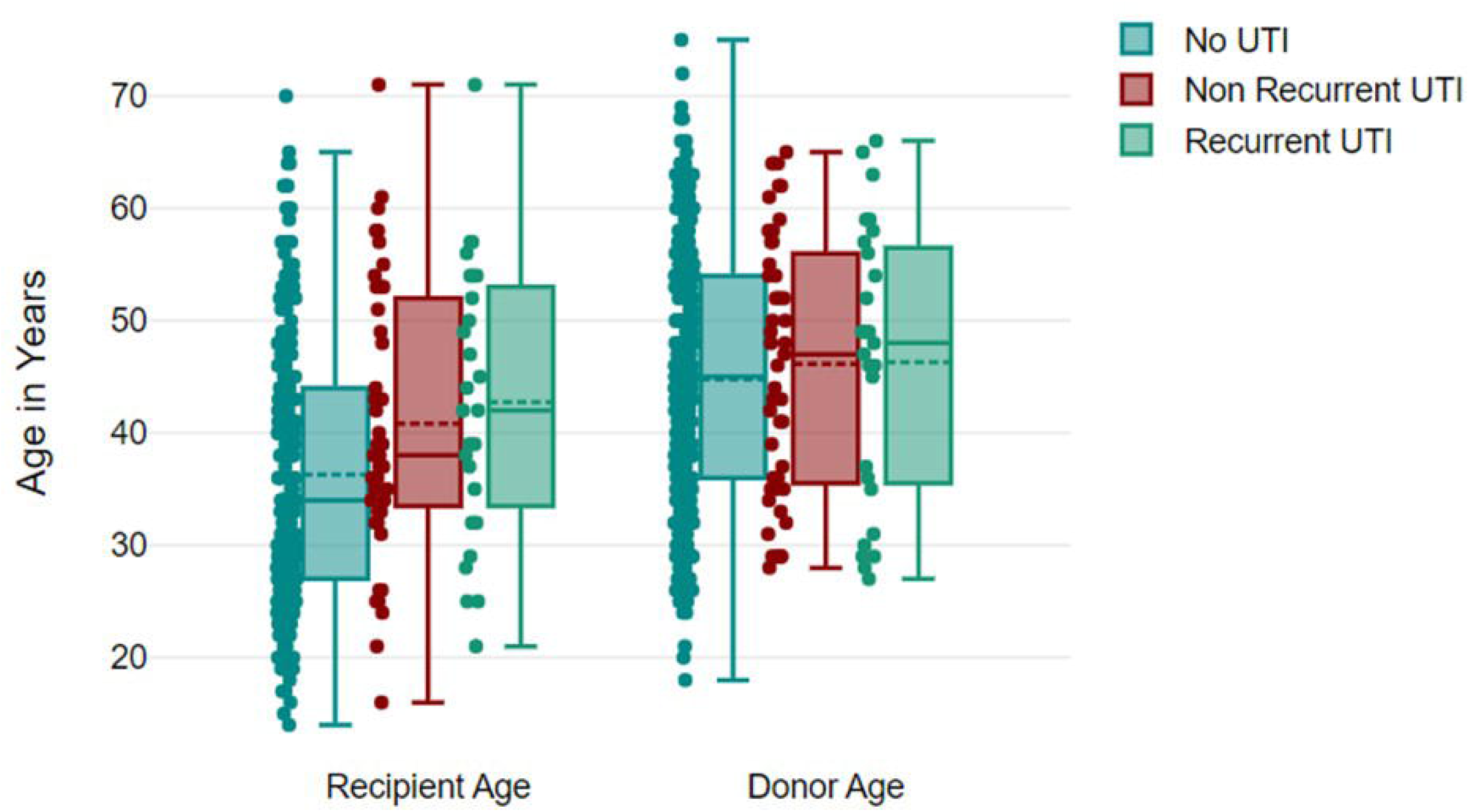
Box and Whisker plot with data points representing the age of renal transplant recipients and donors.

**Figure 4.**
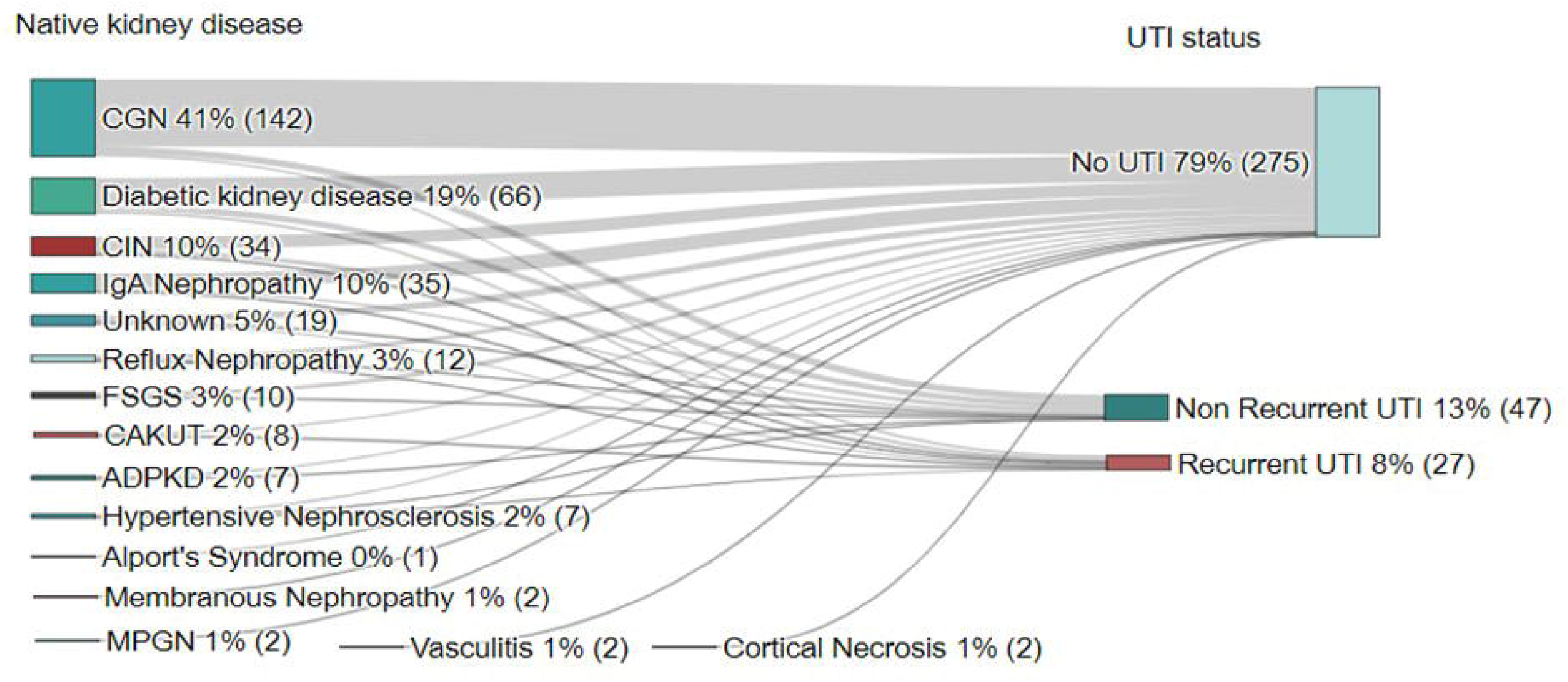
Sankey chart showing the native kidney disease of renal transplant recipients.

### Donor Characteristics

Renal transplant recipients with marginal living donors were associated with R-UTI and NR-UTIs when compared with patients with no episodes of UTI (p=0.007, p=0.003 respectively).

### Transplant Characteristics

Transplant recipients with Post-transplant Diabetes Mellitus (PTDM) were associated with R-UTIs compared with patients with NR-UTIs and patients with No UTI episodes (p=0.005, p=0.002 respectively). Patients who did not receive Trimethoprim-Sulfamethoxazole prophylaxis developed NR-UTIs (vs No UTI, p=0.049). Renal Transplant recipients with delayed graft function had Non-recurrent UTIs (vs. No UTI, p=0.038) but the difference was not statistically significant for Recurrent UTIs (vs. No UTIs, p=0.459). Patients with persistent DJ stents for > 4 weeks had R-UTI (vs No UTI, p < 0.001). Transplant recipients who had not received any induction agents were associated with R-UTI (p=0.019) compared with patients who did not experience any urinary tract infections.

### Number of UTI episodes in each patient

A total of 41(55%) patients experienced only 1 UTI event, 16(22%) patients had 2 UTI events, 10(13%) patients had a sum of 3 UTI events, and 7(10%) patients had more than 3 UTI events (Figure 5).

**Figure 5.**
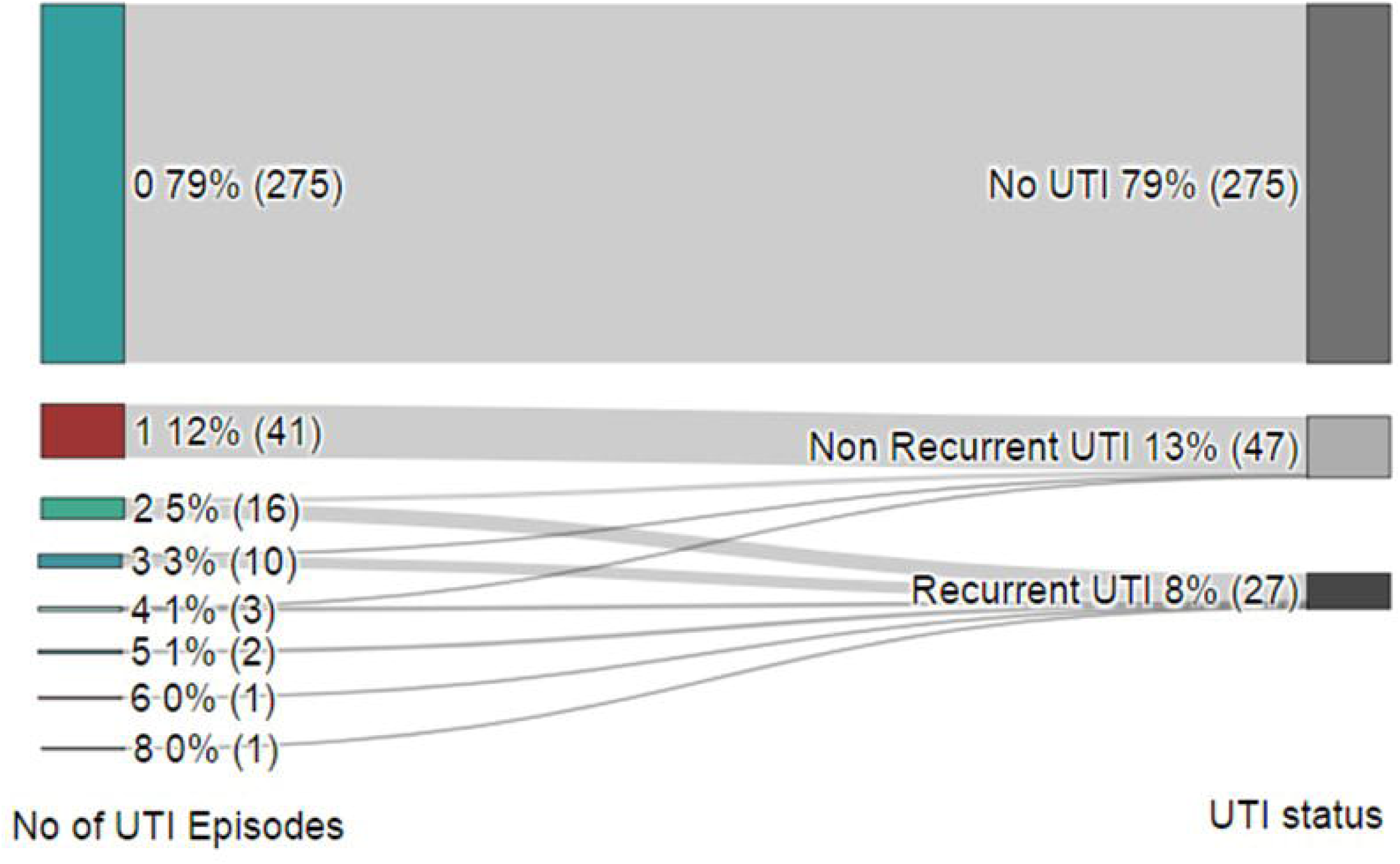
Sankey chart showing the total number of UTI episodes in each patient.

### Median Time for UTI event

The median time to initial UTI in NR-UTI, and R-UTI was 41 days (IQR 16.0 - 624.5 days), and 26 days (IQR 19.5-56.5 days) respectively (Figure 6).

**Figure 6.**
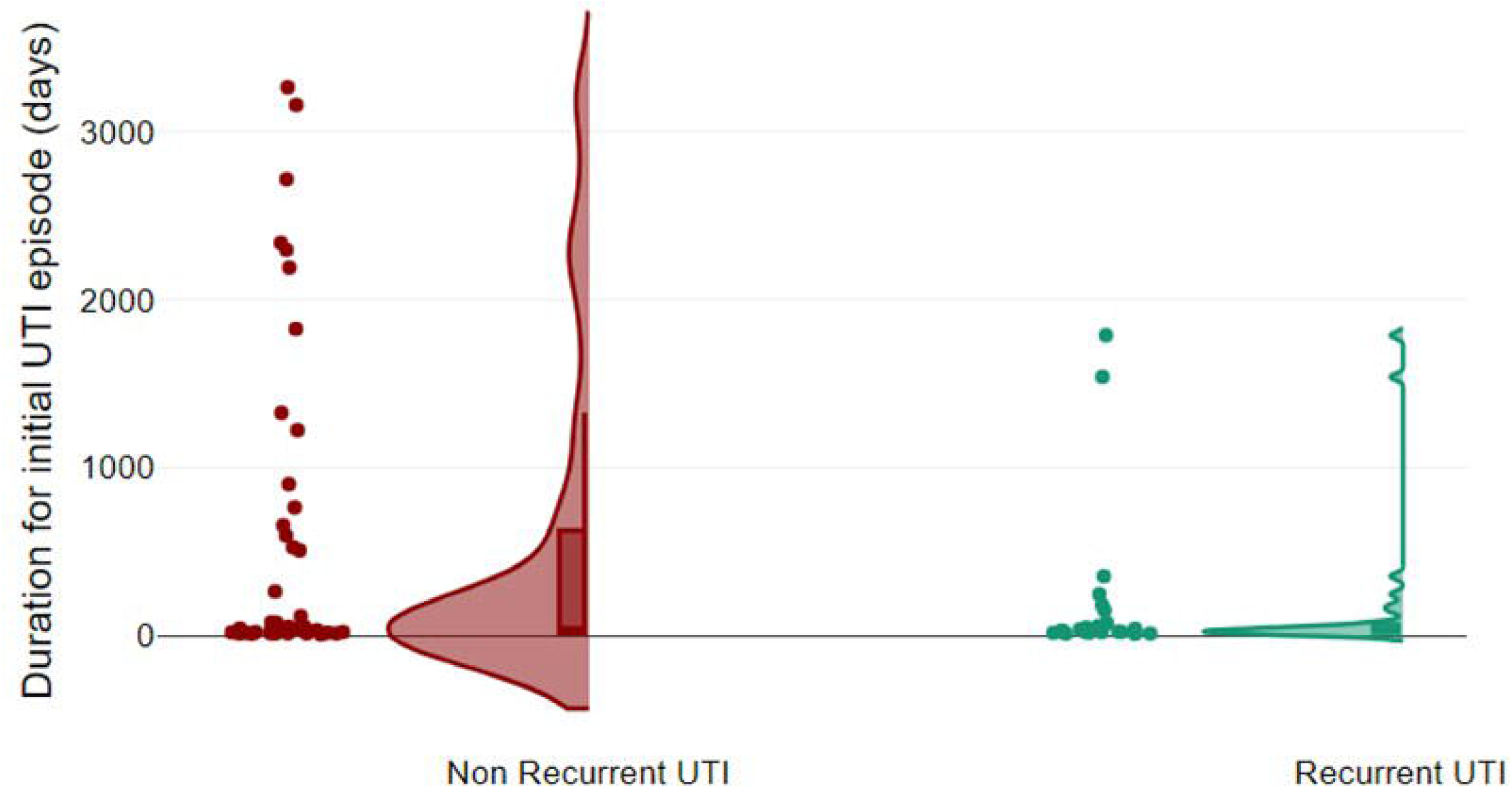
Vertical Raincloud plot showing the duration for the first episode of UTI post-renal transplant in Non-recurrent and Recurrent UTI groups.

### Over-all Graft outcomes

Kaplan-Meier survival analysis showed that post-KT UTI status had a significant effect on graft outcomes (log-rank P=0.002) (Figure 7). Patients experiencing NR-UTI had a significantly greater risk of overall graft failure than those who never developed a UTI [HR 2.312; p = 0.001]. After adjusting for confounding factors in the multivariable Cox regression analysis, NR-UTI [HR 2.318; p=0.001) was associated with a greater risk of graft failure than no UTI or R-UTI (Table 2 and 3).

**Figure 7.**
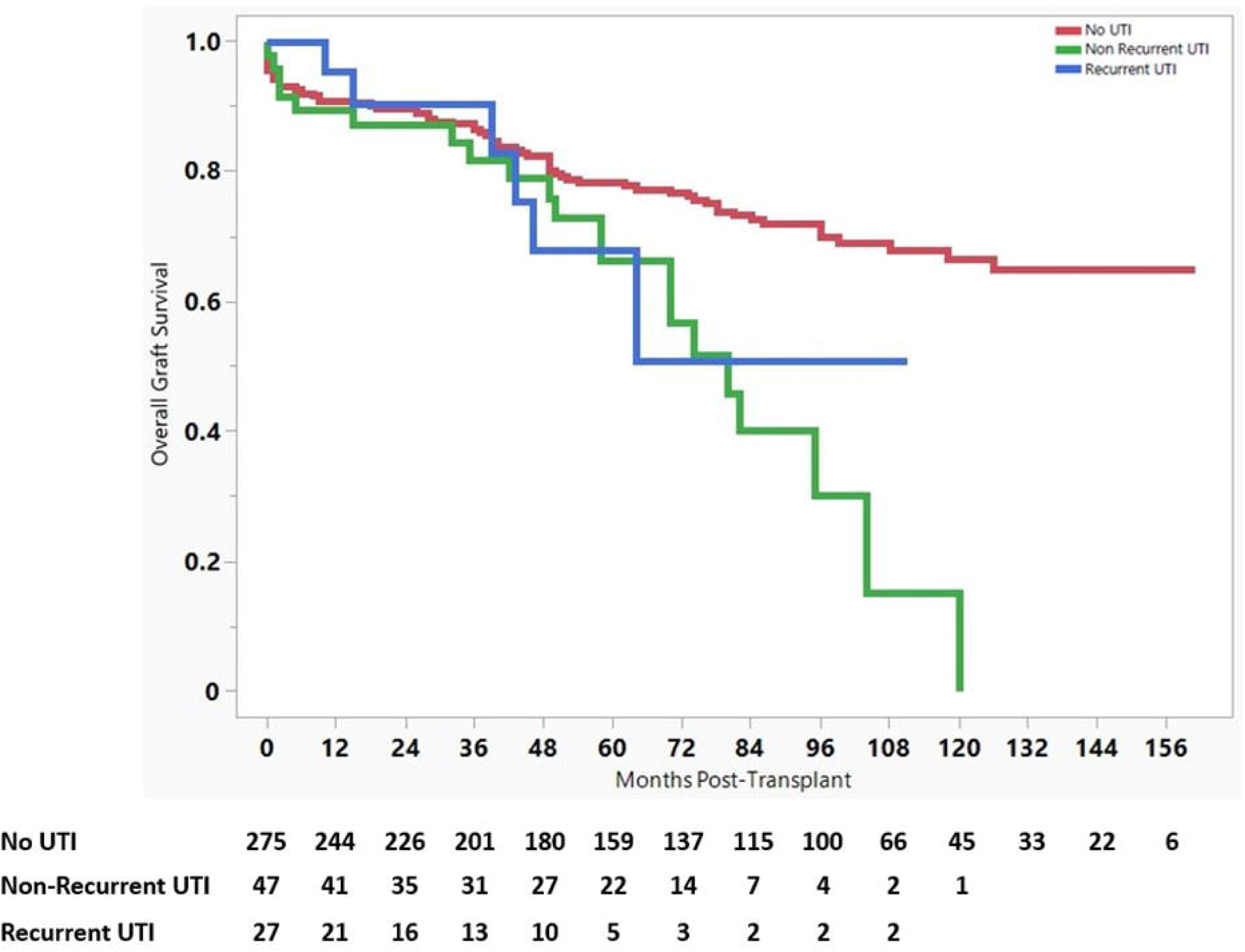
Comparison of overall graft survival by UTI status following renal transplantation. Results are shown for recipients with No UTI, Non-Recurrent UTI, and Recurrent UTI.

**Table 2.**
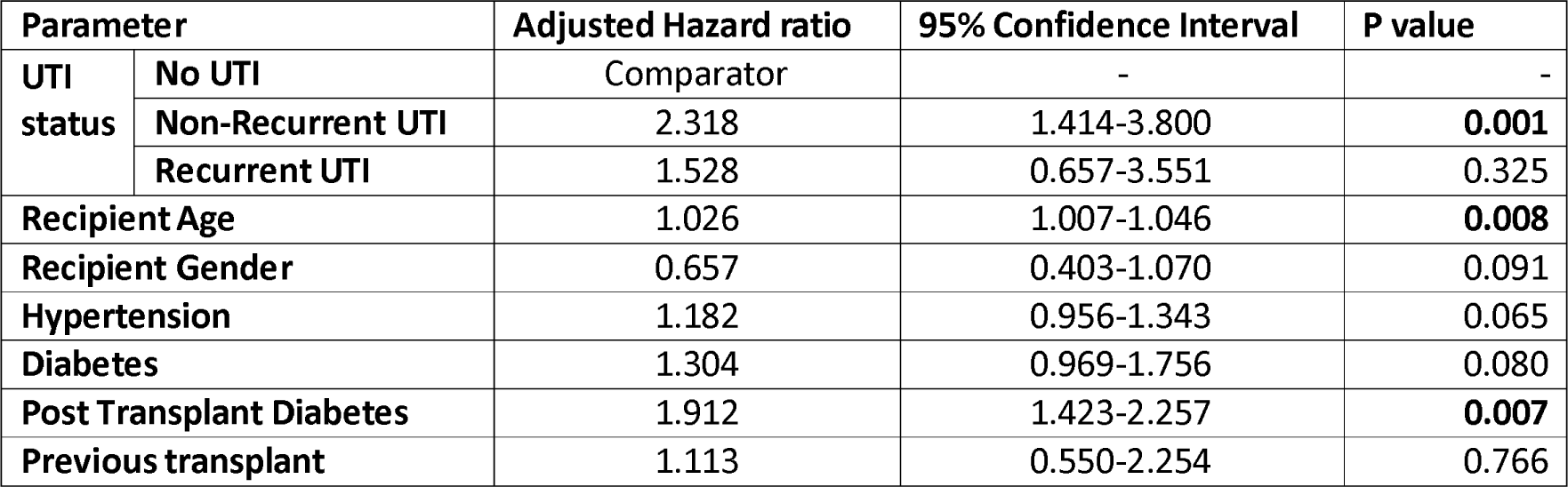
Cox regression model of recipient factors associated with overall graft failure among renal transplant recipients.

**Table 3.**
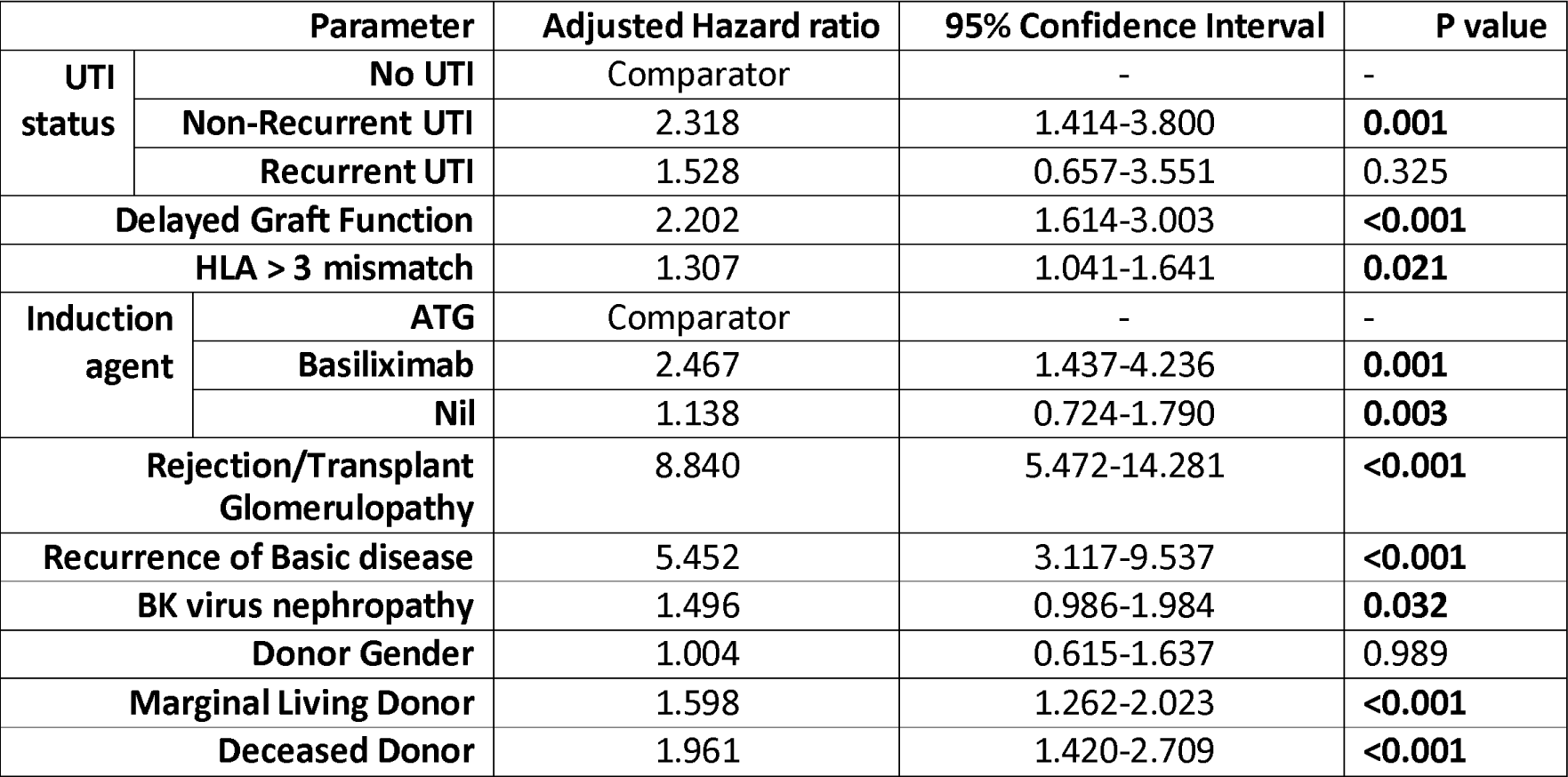
Cox regression model of transplant and donor factors associated with overall graft failure among renal transplant recipients.

| Parameter |  | Adjusted Hazard ratio | 95% Confidence Interval | P value |
| --- | --- | --- | --- | --- |
| UTI status | No UTI | Comparator | - | - |
|  | Non-Recurrent UTI | 2.318 | 1.414-3.800 | **0.001** |
|  | Recurrent UTI | 1.528 | 0.657-3.551 | 0.325 |
| Delayed Graft Function |  | 2.202 | 1.614-3.003 | **<0.001** |
| HLA > 3 mismatch |  | 1.307 | 1.041-1.641 | **0.021** |
| Induction agent | ATG | Comparator | - | - |
|  | Basiliximab | 2.467 | 1.437-4.236 | **0.001** |
|  | Nil | 1.138 | 0.724-1.790 | **0.003** |
| Rejection/Transplant Glomerulopathy |  | 8.840 | 5.472-14.281 | **<0.001** |
| Recurrence of Basic disease |  | 5.452 | 3.117-9.537 | **<0.001** |
| BK virus nephropathy |  | 1.496 | 0.986-1.984 | **0.032** |
| Donor Gender |  | 1.004 | 0.615-1.637 | 0.989 |
| Marginal Living Donor |  | 1.598 | 1.262-2.023 | **<0.001** |
| Deceased Donor |  | 1.961 | 1.420-2.709 | **<0.001** |

A total of 34 (10%) patients had lost graft failure during the observation period, of whom 26 (9%) had no UTI and 8 (17%) had Non-recurrent UTI. The median overall graft survival time was 72 months (95% CI 64.654-79.346). However, the median overall graft survival times for the No-UTI, NR-UTI, and R-UTI groups were 84 months (95% CI 73.763-94.237), 60 months (95% CI 48.817-71.183), and 36 months (95% CI 1.271-70.729) respectively. (Table 4)

**Table 4.** Mean and Median Survival Time of Over-all Graft Survival.

| UTI status | Mean |  |  |  | Median |  |  |  |
| --- | --- | --- | --- | --- | --- | --- | --- | --- |
|  | Estimate | Standard Error | 95% Confidence Interval |  | Estimate | Standard Error | 95% Confidence Interval |  |
|  |  |  | Lower bound | Upper Bound |  |  | Lower Bound | Upper Bound |
| No UTI | 78.032 | 2.730 | 72.680 | 83.383 | 84.000 | 5.223 | 73.763 | 94.237 |
| Non-Recurrent UTI | 61.759 | 5.033 | 51.893 | 71.624 | 60.000 | 5.706 | 48.817 | 71.183 |
| Recurrent UTI | 42.918 | 6.324 | 30.523 | 55.313 | 36.000 | 17.719 | 1.271 | 70.729 |
| Overall | 73.734 | 2.419 | 68.993 | 78.476 | 72.000 | 3.748 | 64.654 | 79.346 |

### Death-censored Graft outcome

Kaplan-Meier survival analysis showed that post-KT UTI status was significantly different in death-censored graft survival (log-rank P=0.024) (Figure 8). Patients experiencing NR-UTI were significantly more likely to experience death-censored graft failure than those who did not experience a UTI (HR 2.365; p = 0.025). The median death-censored graft survival time in the NR-UTI group was 80 months (95% CI 65.627-94.373) (Table 5).

**Figure 8.**
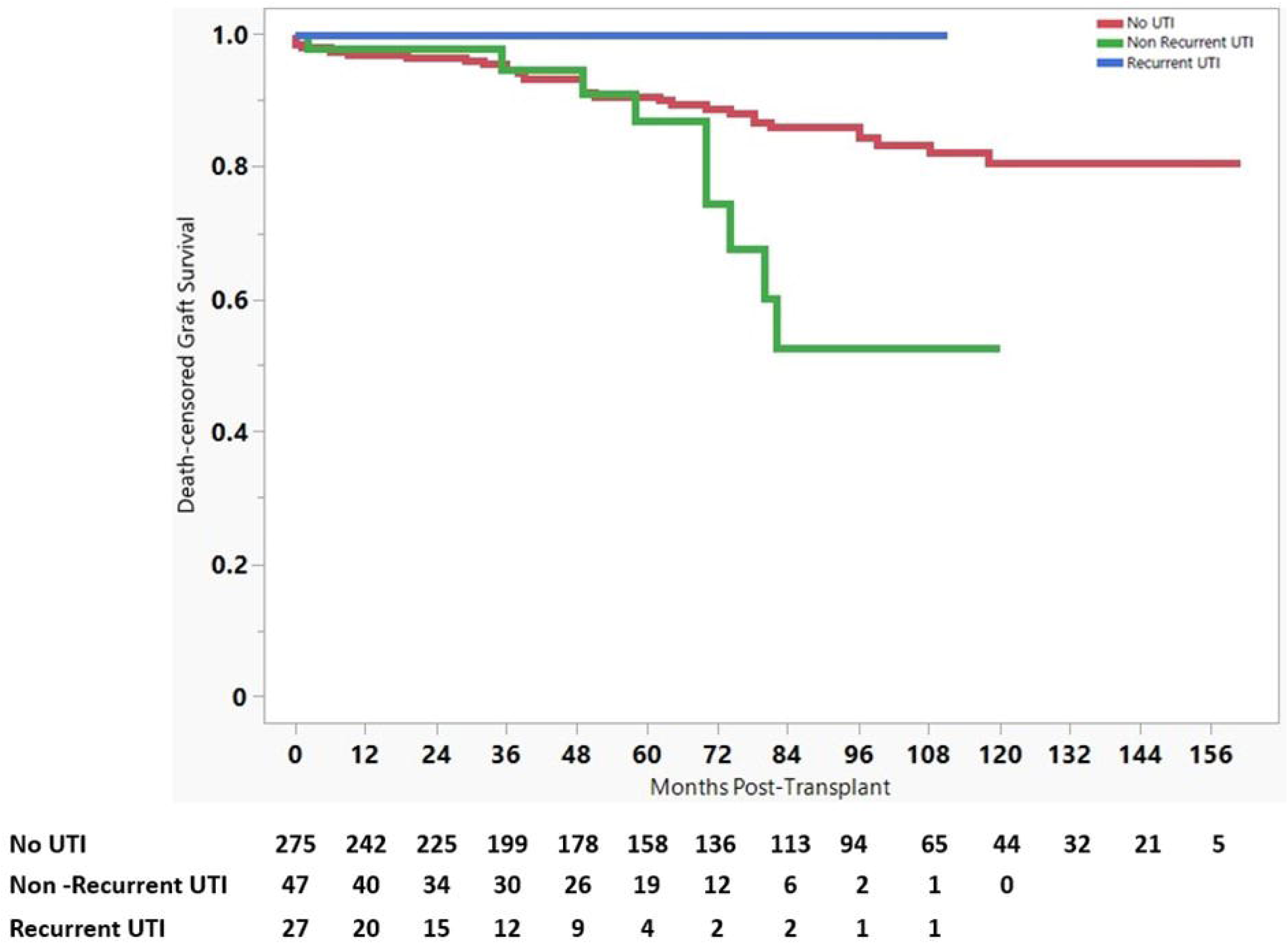
Comparison of death-censored graft survival by UTI status following renal transplantation. Results are shown for recipients with No UTI, Non-Recurrent UTI, and Recurrent UTI.

**Table 5.** Mean and Median Survival Time of Death-censored Graft Survival.

| UTI status | Mean |  |  |  | Median |  |  |  |
| --- | --- | --- | --- | --- | --- | --- | --- | --- |
|  | Estimate | Standard Error | 95% Confidence Interval |  | Estimate | Standard Error | 95% Confidence Interval |  |
|  |  |  | Lower bound | Upper Bound |  |  | Lower Bound | Upper Bound |
| No UTI | 122.086 | 3.872 | 114.497 | 129.675 | - | - | - | - |
| Non-Recurrent UTI | 73.336 | 6.520 | 60.556 | 86.116 | 80.000 | 7.333 | 65.627 | 94.373 |
| Recurrent UTI | 78.168 | 10.511 | 57.566 | 98.771 | - | - | - | - |
| Overall | 116.932 | 3.615 | 109.846 | 124.018 | - | - | - | - |

### Patient outcomes

Kaplan-Meier survival analysis showed that patient survival significantly differed according to post-KT UTI status (log-rank P=0.004) (Figure 9). Patients with NR-UTI and R-UTI had significantly poorer survival rates than those with no UTI. (HR 2.364; P = 0.006, HR 1.517; p = 0.060). A total of 56 (16%) patients died during the observation period, of whom 37 (13%) had No UTI, 13 (27%) had Non-recurrent UTI, and 6 (22%) had Recurrent UTI. The median patient survival time for the NR-UTI group was 118 months (95% CI 90.932-145.068) (Table 6).

**Figure 9.**
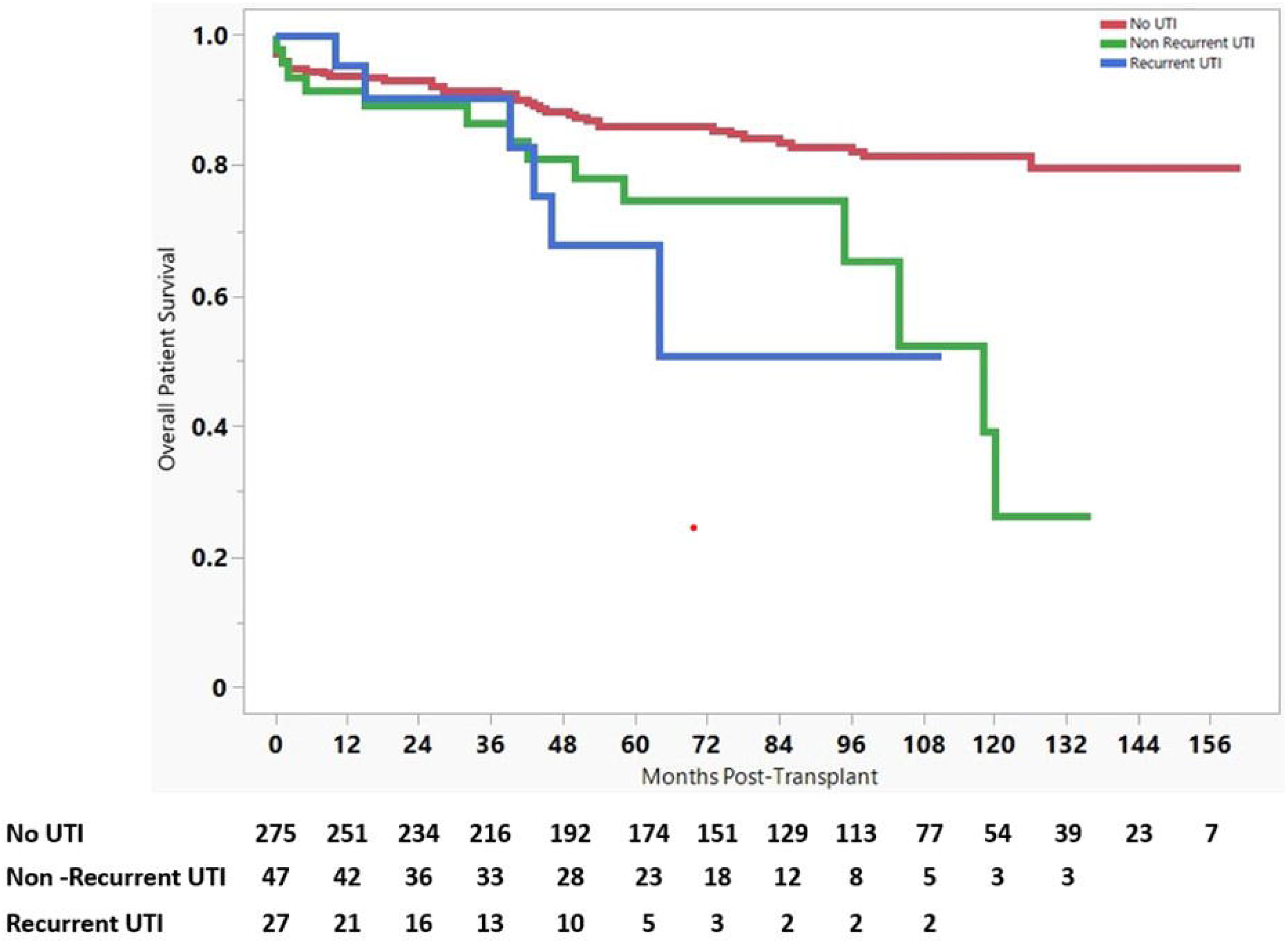
Comparison of overall patient survival by UTI status following renal transplantation. Results are shown for recipients with No UTI, Non-Recurrent UTI, and Recurrent UTI.

**Table 6.** Mean and Median Survival Time of Patient Survival.

| UTI status | Mean |  |  |  | Median |  |  |  |
| --- | --- | --- | --- | --- | --- | --- | --- | --- |
|  | Estimate | Standard Error | 95% Confidence Interval |  | Estimate | Standard Error | 95% Confidence Interval |  |
|  |  |  | Lower bound | Upper Bound |  |  | Lower Bound | Upper Bound |
| No UTI | 137.514 | 3.272 | 131.101 | 143.928 | - | - | - | - |
| Non-Recurrent UTI | 96.168 | 7.748 | 80.982 | 111.353 | 118.000 | 13.810 | 90.932 | 145.068 |
| Recurrent UTI | 78.168 | 10.511 | 57.566 | 98.771 | - | - | - | - |
| Overall | 132.724 | 3.177 | 126.497 | 138.952 | - | - | - | - |

### Microbiological profile and resistance patterns

A total of 140 UTI events were noted in 74 patients during the observation period, of which 133 (95%) were due to Gram-negative bacteria and 7 (5%) were due to Gram-positive bacteria. The most common Gram-negative bacteria were Klebsiella pneumoniae (47%) and Escherichia coli (41%) whereas the most common Gram-positive bacteria were Enterococcus species (Figure 10). Most of the organisms were Multidrug resistant accounting for 38 (73.07%) and 58 (71.6%) events in Non-Recurrent and Recurrent UTIs respectively (Table 7).

**Figure 10.**
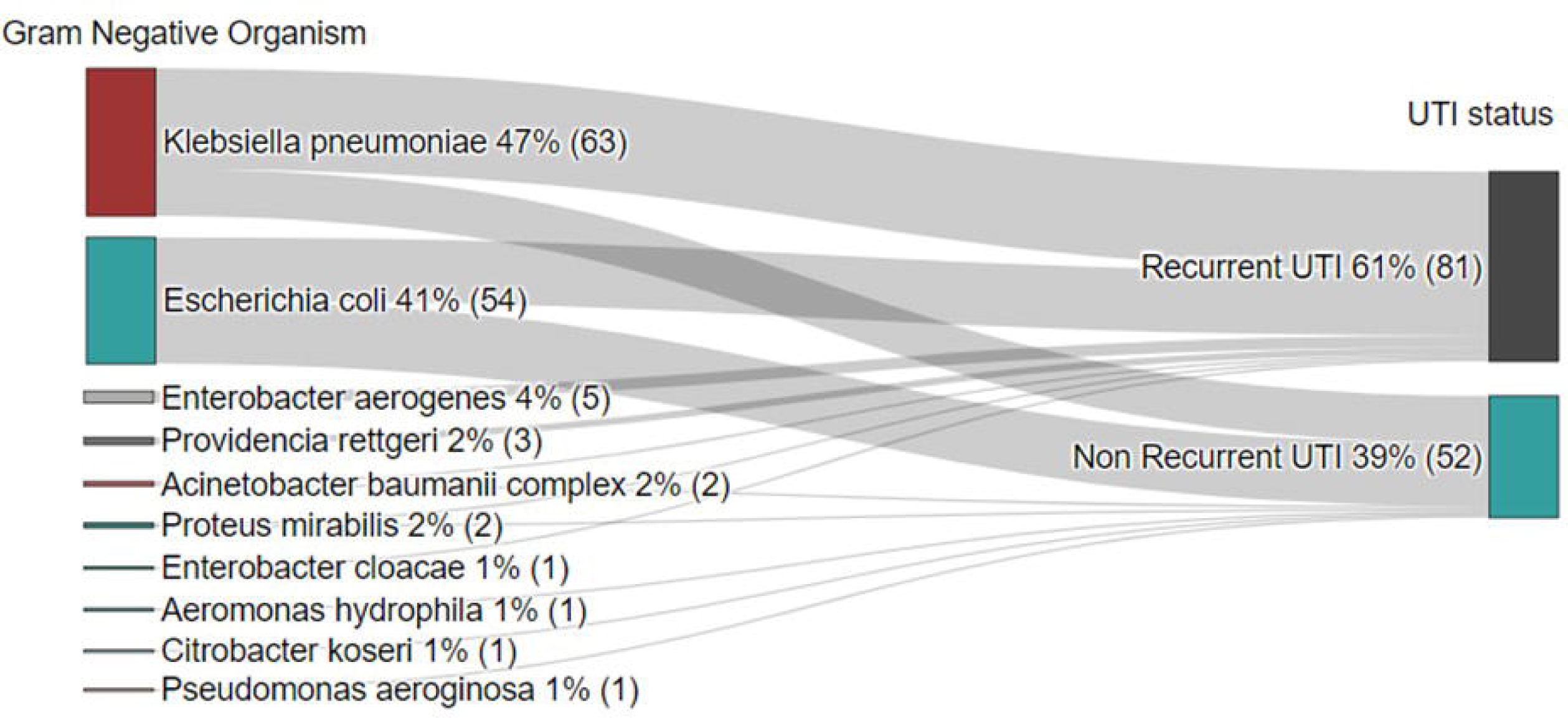
Sankey chart showing the antimicrobial profile of Gram-negative organisms causing post-renal transplant UTI.

**Table 7.**
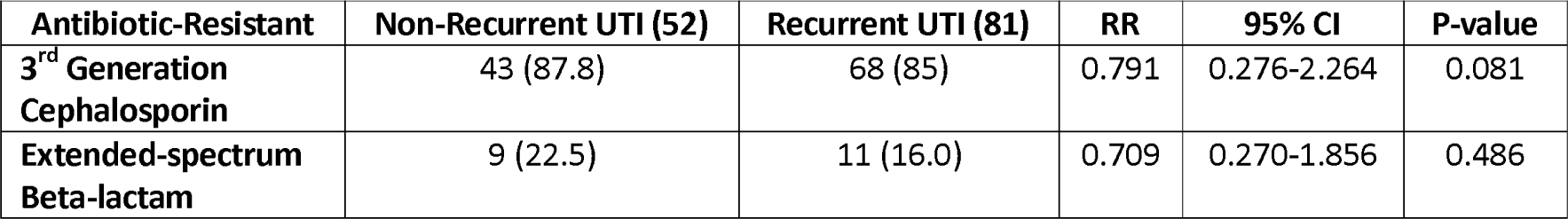
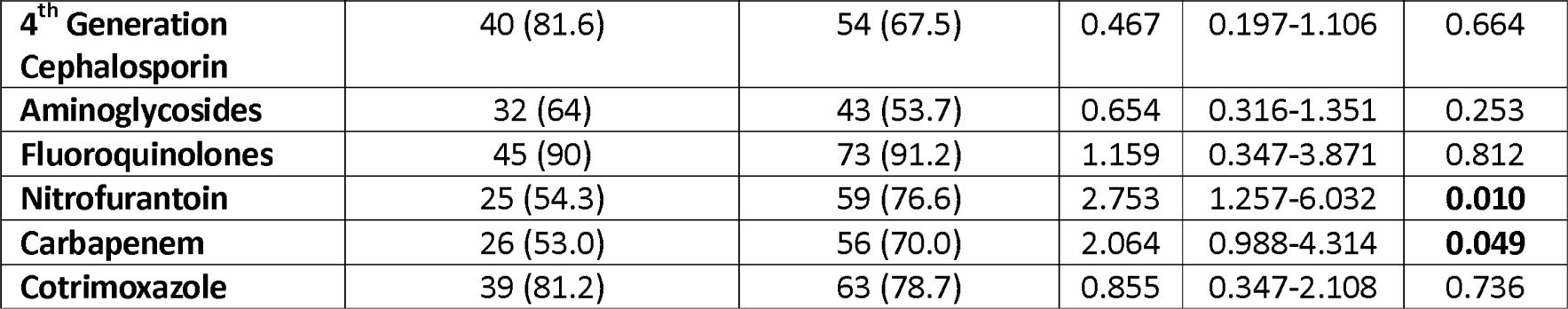
Patient-level analysis of antimicrobial resistance patterns for Gram-negative organisms causing Non-Recurrent and Recurrent post-kidney transplant UTIs.

## Discussion

Although there are no unique criteria for distinguishing early from late UTIs, UTIs occurring within a year after transplantation have been termed early UTIs in numerous studies^1–7^. The timing of UTI episodes post-transplantation is critical, as studies^4–7^ have indicated that early UTI is a risk factor for the development of sepsis and allograft rejection. Similarly, late recurrent UTI increase the likelihood of allograft dysfunction and graft loss^8^. However, these effects have not been consistent across all investigations^4,8,10^. Recent data suggest that even a single episode of UTI can compromise the long-term allograft performance in renal transplant recipients^4,12^.

The processes of allograft damage by pathogens targeting the urinary tract are linked to the UTI-associated inflammatory response to bacterial invasion, which is caused by immunological dysregulation and both local and systemic activation of cytokines such as TNF-α, IL-1, IL-6, and IL-8^7^. Furthermore, cytokine release is thought to play a role in the etiology of allograft rejection by hastening the exposure of allograft tissues to HLAs, resulting in the activation of leukocytes and vascular endothelial cells^7^. In certain situations, the development of acute pyelonephritis can potentially lead to chronic allograft failure due to direct kidney injury^12^.

Infection with virulent Gram-negative uropathogenic organisms with specialized structures such as P fimbriae (pyelonephritis-associated pili) is strongly linked to acute allograft dysfunction^7^. Repeated attacks deplete the regenerative capacity of the graft tissue and promote irreversible fibrosis^7^. As a result, in transplant recipients with recurrent late UTI, allograft damage can result in the formation of numerous localized scarring abnormalities that can be observed by Technetium Tc 99m Di-mercapto succinic acid single photon emission computed tomography (99mTc-DMSA SPECT)^7^.

The present study is unique in that it considered all patients who underwent renal transplantation during the study period, and none were excluded. This study aimed to evaluate and determine the association between UTI after renal transplantation and graft and patient outcomes. In concordance with other studies^4,8^, most cases of UTI events (both R-UTI and NR-UTI) occurred within a year post-transplant. This might be due to the higher net dose of immunosuppression immediately after transplantation.

According to the findings of the current study, older recipients, recipients with marginal living donors, and transplant recipients who had developed DGF or PTDM were more likely to develop UTI. This could be explained by an altered immunological balance, persistently high blood glucose levels, and impaired mucosal barrier function during these special conditions.

A novel finding of the present study was that NR-UTI was associated with a greater risk of graft failure and inferior patient and graft outcomes. This contradicts most previous studies^4,8^ that showed that R-UTIs are generally associated with worse patient and graft outcomes. Although the current study demonstrated an increase in mortality in R-UTI patients, it was not statistically significant when compared with those with No UTI. A possible explanation for the lower graft survival among NR-UTI compared to R-UTI could be that patients with R-UTI received empirical treatment with carbapenems, prolonged antibiotic prophylaxis, close monitoring and follow-up. This could be due to the intensive approach to root cause analysis of Recurrent UTI and early treatment strategies. Some patients had reflux to the native kidney and underwent surgical interventional procedures (1 patient underwent native kidney nephrectomy, and 3 transplant recipients had Dextranomer/hyaluronic acid bulking agent injected at the vesicoureteral junction). However, the discrepancy may be attributable to the shorter follow-up time in these instances, as most patients with R-UTIs had undergone transplants recently (within the last 5 years).

India, is a large and diversified country, with a wide range of infectious disease concerns owing to population density, inadequate sanitation, economic inequality, access to healthcare, and variable levels of vaccine coverage. Due to the increased morbidity and mortality, it is connected with, anti-microbial resistance has become a serious health concern. Similar to previous studies, Gram-negative organisms accounted for the majority of UTIs, with Klebsiella pneumoniae and Escherichia coli contributing approximately 47% and 41%, respectively. The pathogens causing R-UTIs were analogous to those implicated in NR-UTI, with no statistically significant differences observed between the two groups for any organism.

When analyzed at the patient and UTI event levels, both NR-UTI and R-UTIs were more likely to be caused by multidrug-resistant organisms. Resistance to Carbapenems or Nitrofurantoin is associated with the development of Recurrent UTIs, leading to treatment challenges. This is probably due to the ineffective practice of antibiotic stewardship, and thus there is an increase in multidrug-resistant pathogens especially Nitrofurantoin and Carbapenem resistance, as these antibiotics are currently regularly used to treat complicated UTIs.

The divergent outcomes observed in the current study could be explained by the early treatment techniques and the root cause analysis approach used in the evaluation of R-UTIs. The current study’s significant drawback is that the majority of R-UTI patients underwent recent transplants; therefore, their long-term graft (>5 years) outcome could not be studied.

The inclusion of all kidney transplant recipients throughout the research period and the extensive follow-up period were strengths of this study. An independent risk factor evaluation of UTI-related graft dysfunction was performed after the influence of confounding variables was assessed using a multivariable Cox proportional regression analysis. To quantify the independent risk of confounding factors, proportional hazards, and Hazard ratios were obtained. Asymptomatic bacteriuria if present in the initial 2 months post-transplant, was reconfirmed with a repeat culture and treated if the same organism was isolated^14^.

However, this study has several limitations. We did not assess the overall prevalence of asymptomatic bacteriuria and its relationship with the emergence of UTI following renal transplantation. As this was a single-center study, it was not possible to extrapolate the findings to the entire target population. The current study included female patients who were underrepresented as transplant recipients and strongly overrepresented donors. This could be due to the predominance of donors in India being either mother or spouse^15^. Hence the results cannot be generalized to other transplant centers with a balanced sex distribution of recipients and donors. This study did not explore the relationship between various therapy types and durations and their impact on the results.

In summary, we discovered that the NR-UTI group had consistently declining graft performance, poorer graft survival, and poorer patient outcomes than those in the no-UTI group. The most likely time for UTIs following a kidney transplant is within one year after the transplant, and the most prevalent cause is multidrug-resistant Gram-negative bacteria. Additionally, it was discovered that certain factors were independently associated with graft failure, including male recipients, post-transplant diabetes mellitus, transplant glomerulopathy, recurrence of basic disease/de-novo glomerular disease, marginal living donors, deceased donors, delayed graft function, HLA > 3 mismatches, BK virus nephropathy, rejection episodes, nil induction, and basiliximab used as an induction agent.

## Conclusions

Given the scarcity of available treatments, it is vital to combat multi-drug antimicrobial resistance, particularly carbapenem resistance. In individuals who have had kidney transplantation, even a single UTI can result in poor long-term graft and patient outcomes.

## Compliance with Ethical Standards

### Conflict of interest

The authors have declared that no conflict of interest exists.

### Ethical statement

This study was conducted following the principles of the Declaration of Helsinki. The original study protocol was approved by the Institutional Ethics Committee of Ramaiah Medical College on 22-02-2023, its Institutional Review Board (DRP/IFP1085/2023).

### Informed consent

Waiver for informed consent was approved by Institutional Review Board

## Data Availability

All data produced in the present study are available upon reasonable request to the authors

## Acknowledgments

Not received any financial grant for the study

## Notes

### Competing Interest Statement

The authors have declared no competing interest.

### Funding Statement

This study did not receive any funding

### Author Declarations

Ethics Committee of Ramaiah Medical College, its Institutional Review Board had received the study protocol and meeting was conducted and gave ethical approval for the study on 22-02-2023 (DRP/IFP1085/2023). As the study is retrospective, waiver for consent was provided by IRB

